# Dimensions, social and healthcare setting determinants of client satisfaction for HIV/AIDS services: a systematic review

**DOI:** 10.1101/2023.07.10.23292462

**Authors:** Aklilu Endalamaw, Charles F Gilks, Fentie Ambaw, Mark D. Chatfield, Yibeltal Assefa

## Abstract

**Introduction:** Quality HIV/AIDS care affects clients’ health-seeking behaviour and adherence to services, which can be evaluated through the patient’s satisfaction with the services. There was an information gap on the status and contributors to HIV/AIDS clients satisfaction, which supports comprehending universal health coverage. This review presented dimensions and comprehensive determinants of HIV/AIDS clients satisfaction.

**Methods:** We conducted a systematic review based on articles from Web of Science, EMBASE, PubMed, Google Scholar, and grey literature sources. Studies that have reported satisfaction of people living with HIV or at least one determinant and are written in English with full-text available were included. Findings from each study were described in a narrative description.

**Results:** There was a heterogeneous level of HIV/AIDS clients’ satisfaction in different settings and countries. Accessibility of services, confidentiality, waiting time to receive care, technical competency, knowledge, and attitude of health care workers were frequently reported determinants. Educational and employment status were common modifiable social factors.

**Conclusions:** Public health programmes should follow a multi-pronged approach to fulfil people living with HIV clients’ healthcare needs. It is vital to improve HIV care integration with primary healthcare, improve financing of HIV care, digitalise healthcare settings, invest in pre-service education and in-service training, provide culturally sensitive services, strengthen social works and behavioural change communication services.

## Introduction

HIV/AIDS has remained an epidemic disease, causing many countries in the world to face health, economic, and societal challenges due to the large number of people living with HIV (1). Different strategies, such as “90-90-90” and “goals of ending the HIV epidemic”, are endorsed to halt HIV transmission (2). These strategies need to be implemented while maintaining a minimum of acceptable healthcare standards, ‘leaving no one behind’ quality of care (3, 4).

Therefore, quality of care has been given due attention (5), which can be evaluated by using patient satisfaction (6). Satisfied people who live with HIV are more likely to have good antiretroviral therapy adherence and not lose their regular healthcare schedule (7). Measuring clients’ satisfaction discloses the effectiveness of healthcare provision and the performance of healthcare workers (8, 9). It also ensures whether healthcare strategies are implemented effectively with the standard of care and have an impact on ending the HIV epidemic (10) which should be evaluated continuously.

Patient satisfaction involves collating feedback using different tools and domains (11, 12). Domains include client-patient interaction, service accessibility, therapeutic communication, timely care, culturally sensitive care, confidentiality, and privacy (13, 14). These domains reflect how patient-centred care should be. The World Health Organisation cherishes the significance of patient satisfaction in improving healthcare services everywhere in the world. These domains can be assessed using medical record reviews, direct patient feedback mechanisms, technology assisted devices (online platforms), and interviews (15–17). However, there was no comprehensive evidence about the clients’ satisfaction with HIV/AIDS services despite original studies in different countries (18–23).

Accordingly, the purpose of this review was to synthesis HIV/AIDS clients satisfaction and identify determinants of the healthcare services they received. The findings from the review will allow HIV programme planners and clinicians to make evidence-based and informed decisions to improve the quality of HIV care services.

## Methods

### Protocol registration and reporting

The ‘Preferred Reporting Items for Systematic Reviews and Meta-analysis’ guideline was used to conduct and report the review findings (24). The topic was registered in ‘the International Prospective Register of Systematic Reviews’ with registration number CRD42020207215.

### Information sources and search

PubMed, EMBASE, and Web of Science core electronic databases were searched. We also manually searched Google Scholar and cross-references. The last search was on 12 June 2021.

The search included any combination of the following search terms: ‘human immunodeficiency virus’, ‘acquired immunodeficiency syndrome’, “tuberculosis”, “TB”, “HIV”, “AIDS”, “patient”, “satisfaction”, “determinants”, “predictors”, “risk factors”, “associated factors”, and “factors”. The Boolean operators “AND”/“OR” conjunctions were used to combine terms. An example search strategy fitted for PubMed was (((((“HIV” [MeSH Terms] OR HIV OR “Acquired immunodeficiency syndrome” OR AIDS OR “Human immunodeficiency virus/Acquired immunodeficiency syndrome” OR HIV/AIDS) OR ((Tuberculosis [MeSH Terms] OR TB OR tuberculosis))) OR (((((“Tuberculosis“[Mesh]) AND (“HIV“[Mesh])) OR ((“Tuberculosis“[Mesh]) AND (“Coinfection“[Mesh]))) OR ((“HIV“[Mesh]) AND (“Coinfection“[Mesh]))) OR (((((HIV[tiab]) OR (“human immunodeficiency virus“[tiab])) AND ((co-infection[tiab]) OR (coinfection[tiab]))) OR (((TB[tiab]) OR (tuberculosis[tiab])) AND ((co-infection[tiab]) OR (coinfection[tiab])))) OR (((TB[tiab]) OR (tuberculosis[tiab])) AND ((HIV[tiab]) OR (“human immunodeficieny virus“[tiab])))))) AND ((“Patient satisfaction” [MeSH Terms]) OR “patient experience” OR “patient priorities” OR “client satisfaction” OR “user satisfaction“)) AND (determinants OR predictors OR “factors affecting” OR measurement OR dimensions OR aspects OR attributes OR “associated factors” OR “risk factors“)) NOT ((“Review” [Publication Type]) OR (“Clinical Conference” [Publication Type])).

### Eligibility criteria, study selection and data extraction

Inclusion criteria were those studies report the satisfaction or at least one determinant for people living with HIV/AIDS services conducted amongst all age groups and published in English with full-text available. These services include HIV services (ART and opportunistic infections treatments, clinical consultation and preventive, rehabilitative, and palliative care services), pharmaceutical services and ART laboratory services.

We did not restrict the date of publication and country. When we found two or more studies with the same contents, studies published first if have complete data were included. We excluded studies conducted other than the English, qualitative studies, conference presentation, and citation and abstract only.

All citations were exported to the EndNote X7 reference library to remove duplicates (25). Screening was made according to the inclusion and exclusion criteria. First, title screening was performed. Second, articles were evaluated for abstract. Then, their full texts were evaluated to be included in the review.

Articles eligible for full-text review were extracted using a data extraction form using Microsoft Excel 2010. Data were collected about characteristics of the articles include data collection period, country, study population, study design, satisfaction measurement tool, data collection method, sample size, and main findings. In extracting the main findings, the satisfaction level was dependent variable, while individual, health worker and healthcare setting were independent variables.

### Risk of bias assessment

The “Newcastle-Ottawa Scale” was applied to evaluate the quality of each study (26). The study considered as low risk for bias when it scores ≥50% and above the value of assessment criteria, and high risk for bias when the score of each assessment criteria became below the 50% of criteria (Supplementary file 2).

### Data analysis

Studies that have appeared with reported satisfaction levels using proportion, mean value or specific dimensions or its associated factors were systematically described. Determinants were grouped in the personal, health worker, and healthcare settings levels. Results were presented by figures and tables. The pooled percentage and trend over time of satisfied HIV clients was not performed because of heterogeneity of measurement tool in the included articles. Additionally, sensitivity analysis, regression analysis and publication bias did not be performed.

## Results

### Search results

The search process accessed 4,848 articles. Of these, 58 were from EMBASE, 164 were from Google Scholar, 1,497 were from PubMed and 3,129 were from Web of Science. A total of 4,720 articles remained after duplicates removed. Then, 4,565 articles were excluded because of unrelated titles and citations only. Of the articles that got the full-text screening phase, 33 were excluded. To end with, 66 articles included in the final review (Figure 1); 32 were available in Web of Sciences and PubMed, 11 in PubMed, 17 in Google Scholar, 3 in all PubMed, Web of Sciences and EMBASE, 1 in both PubMed and EMBASE, and 2 grey literatures from Google.

**Figure 1:**
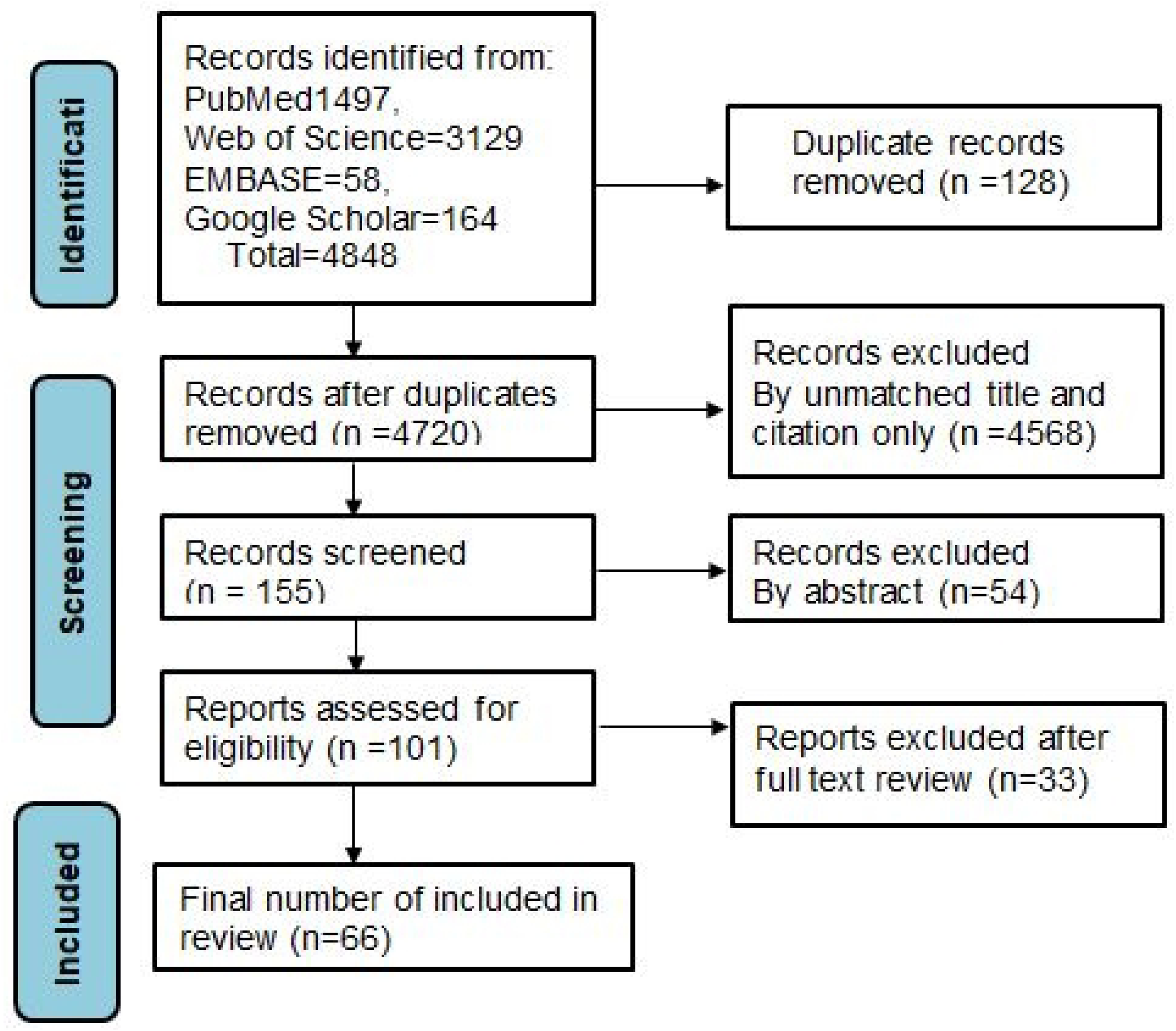
Article selection process

### Characteristics of articles

The final review has included a total of 66 studies. Authors collected data between 1988 (27) and 2019 (28), and published it between 1993 and 2021. According to the WHO region, 42 articles were published in Africa (19, 22, 23, 28–66), 9 studies in the South-East Asia (18, 67–74), 9 studies in the America (7, 27, 75–81), 4 in the Western Pacific (20, 82–84) and 2 in the European (21, 85) region. Specific to country, 17 were from Ethiopia, 12 from Nigeria, 7 from India, 6 each from South Africa and the United States of America, 3 from Tanzania, 2 each from Brazil, Vietnam and Zambia, and 1 each from United Kingdom and Kenya, Australia, Cameroon, Canada, China, France, Kenya, Russia, and Thailand. Regarding the study population, two among paediatrics and caregivers (28, 49) and one among all age groups (37), two did not report the age category (27, 81) and the remaining 61 were conducted among the adult age group. One study was conducted among males (80) and two of the studies were among women (76, 78) while others were among both sexes (Table 1).

**Table1:**
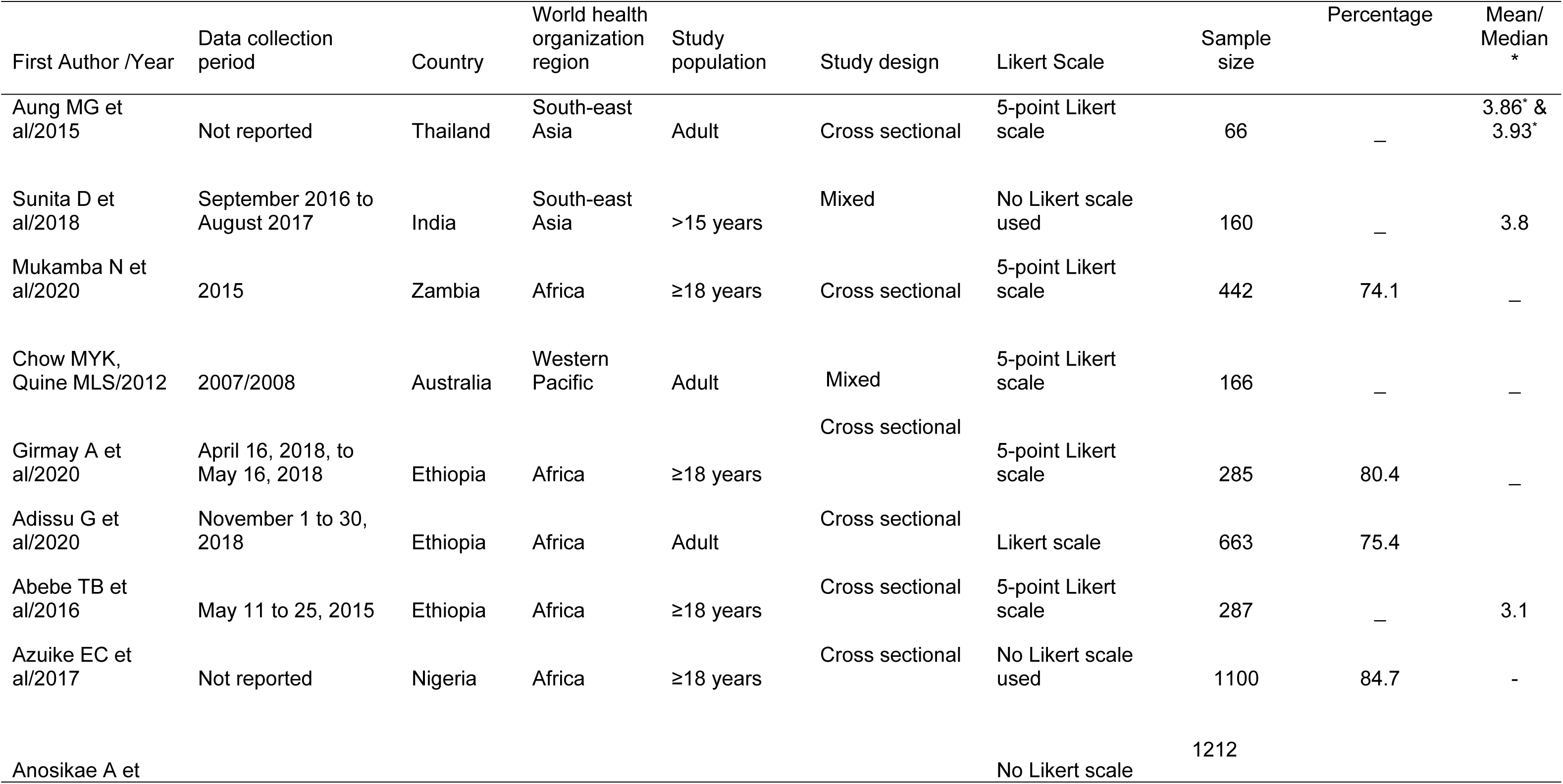
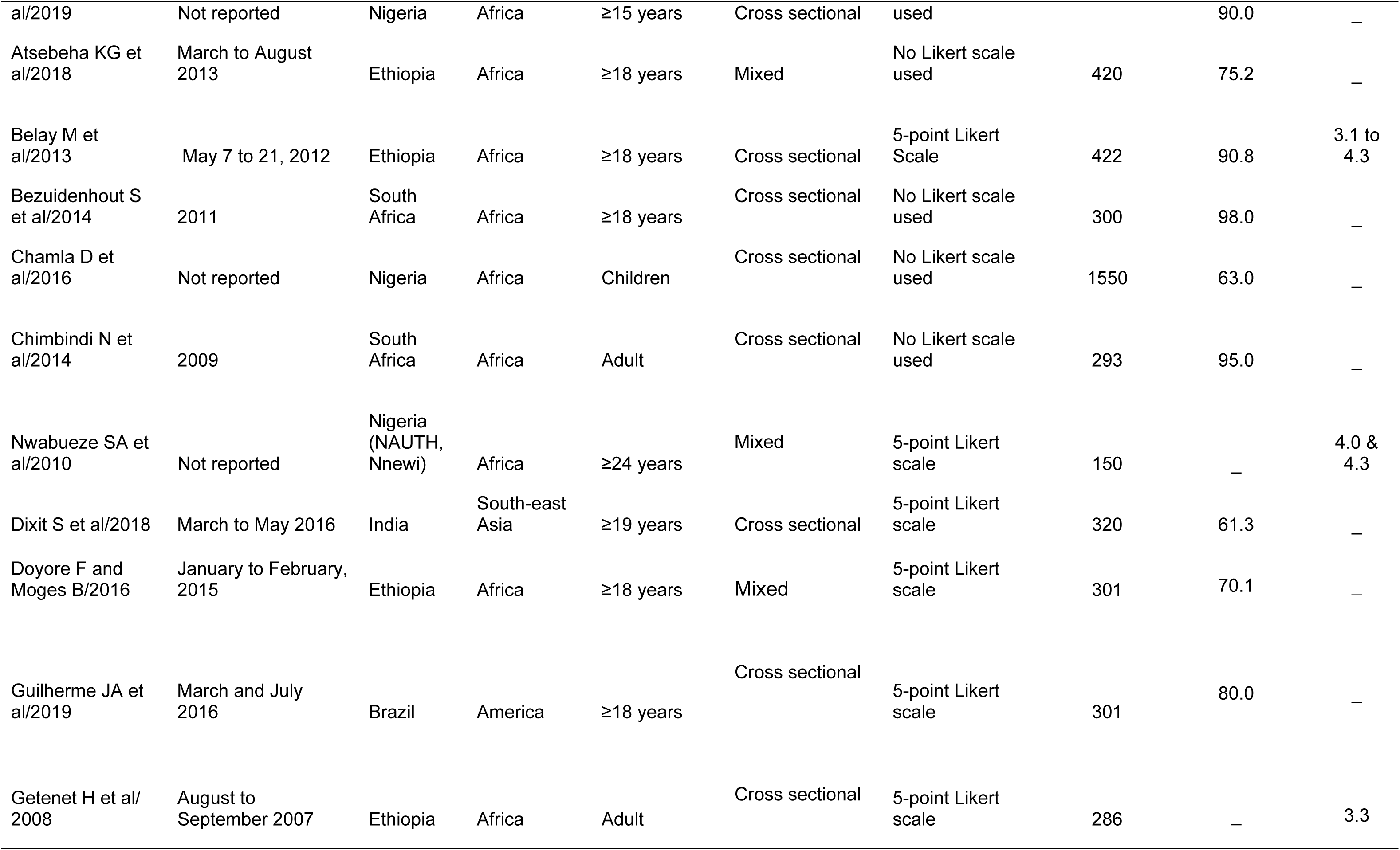
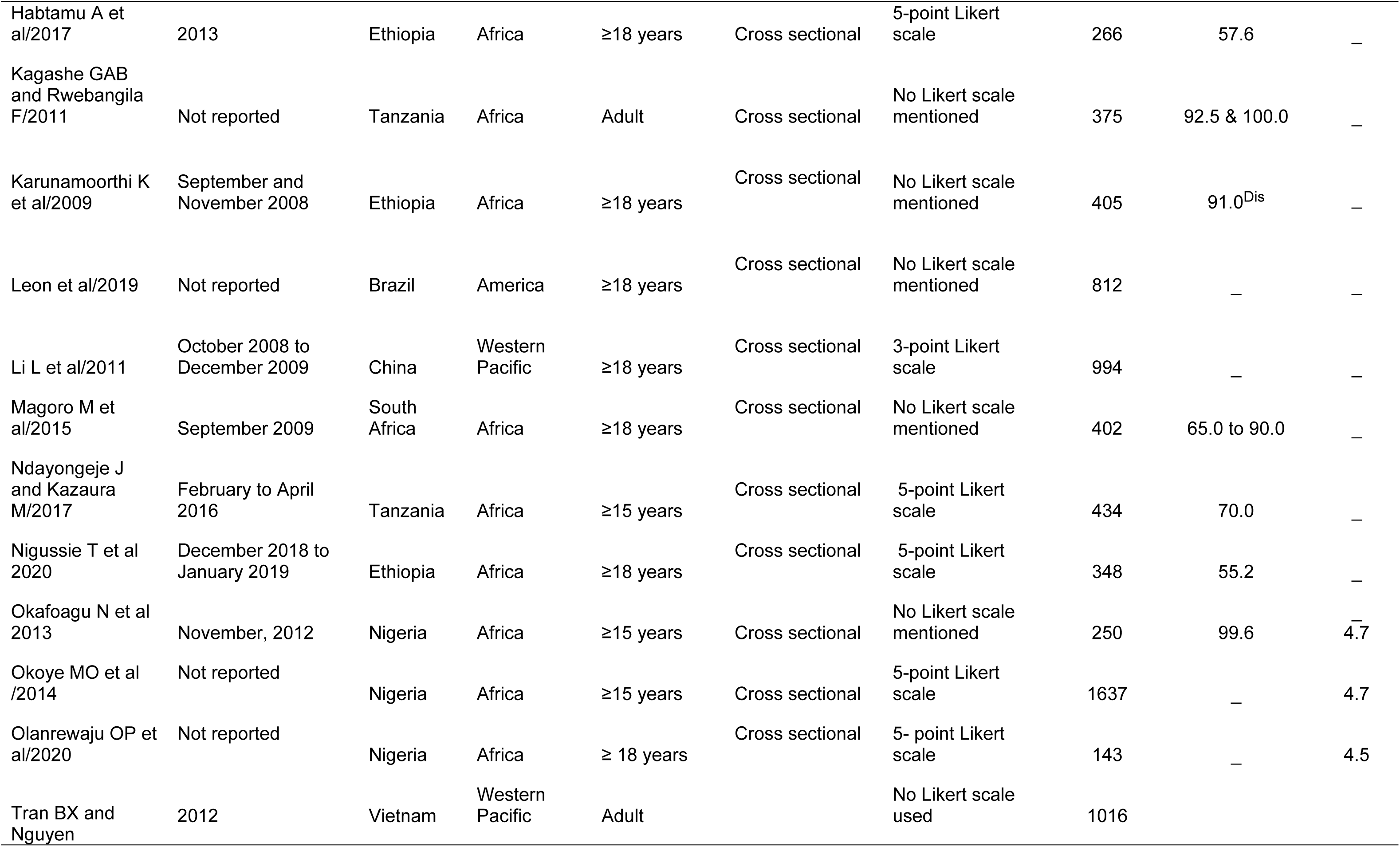
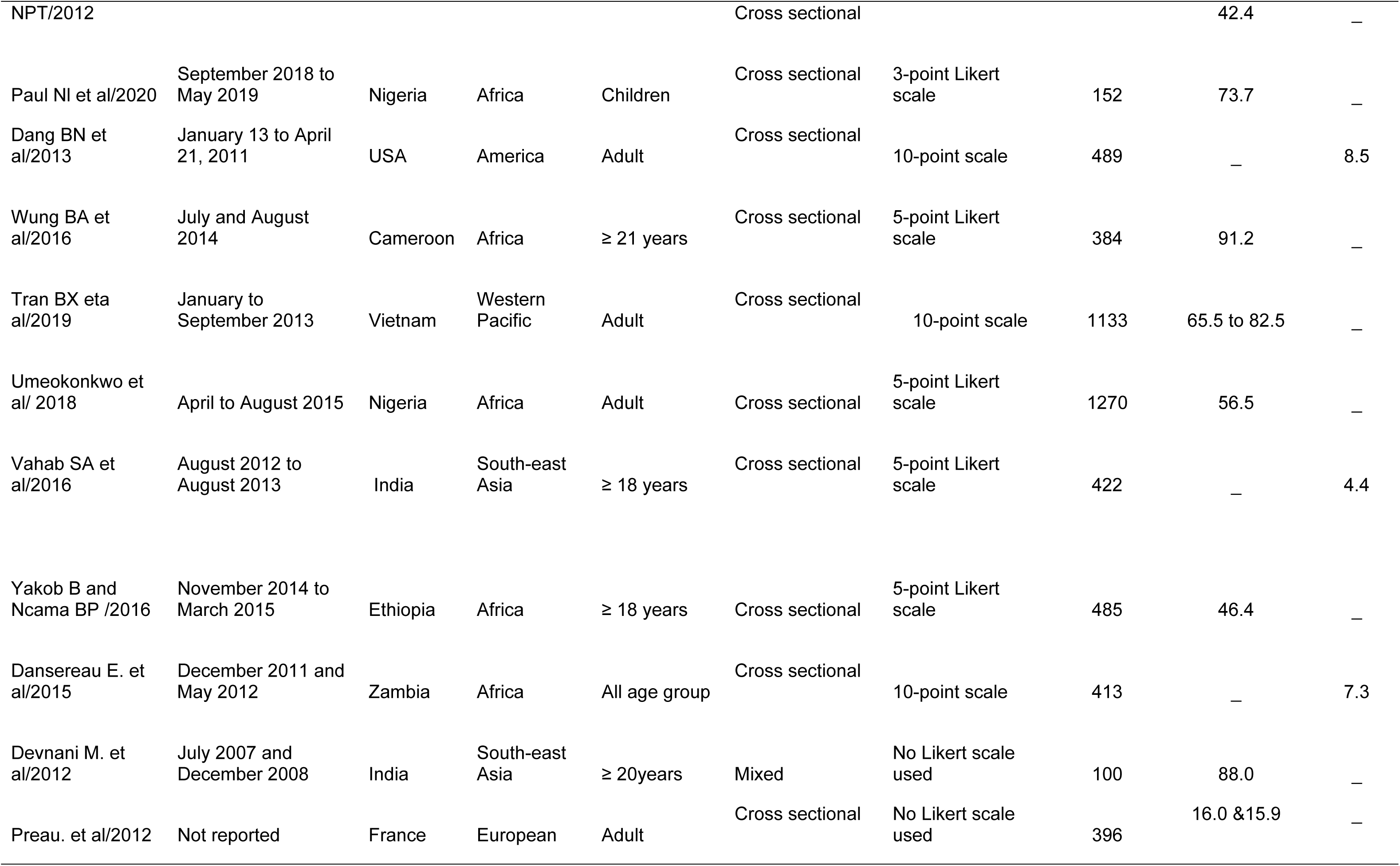
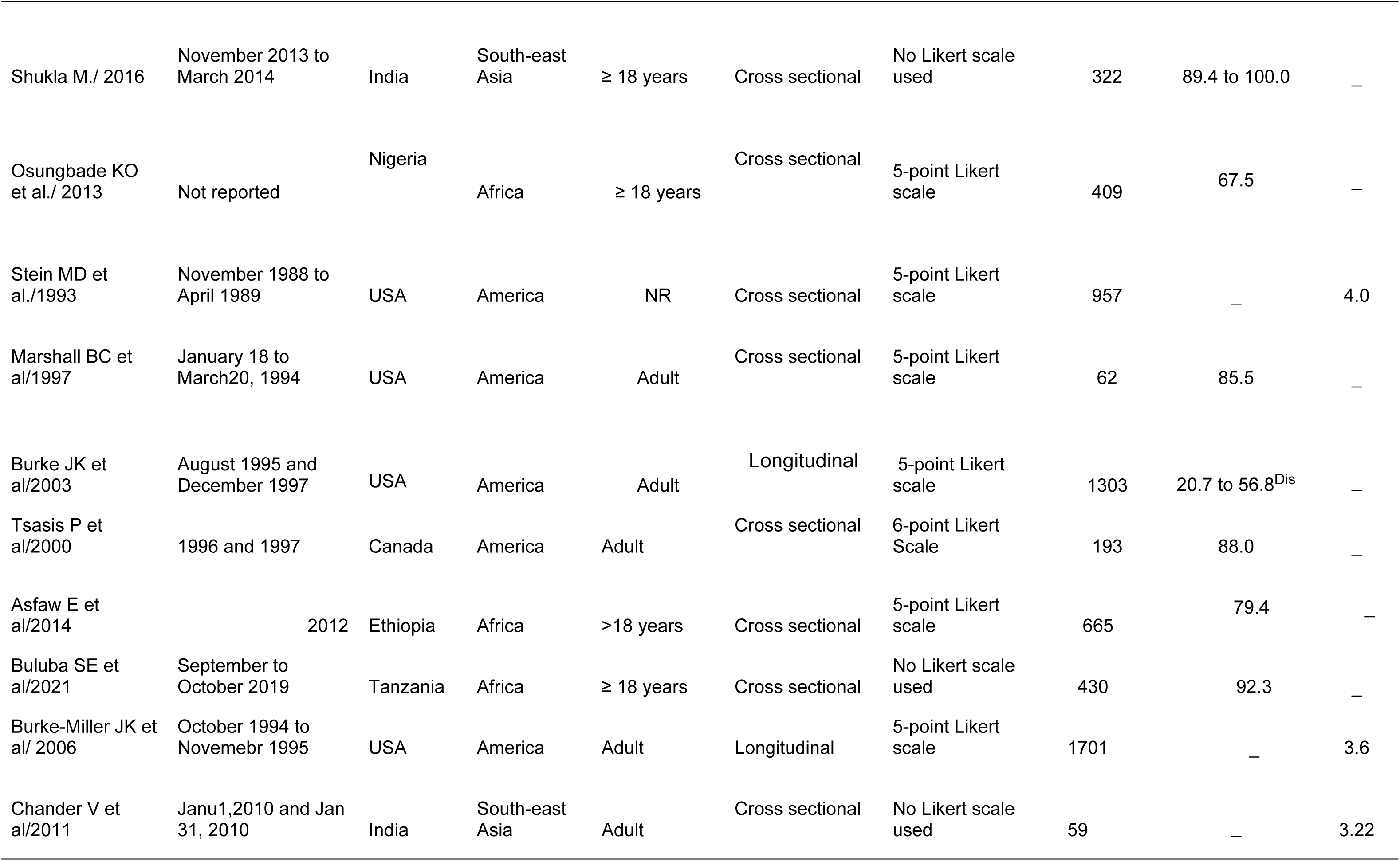
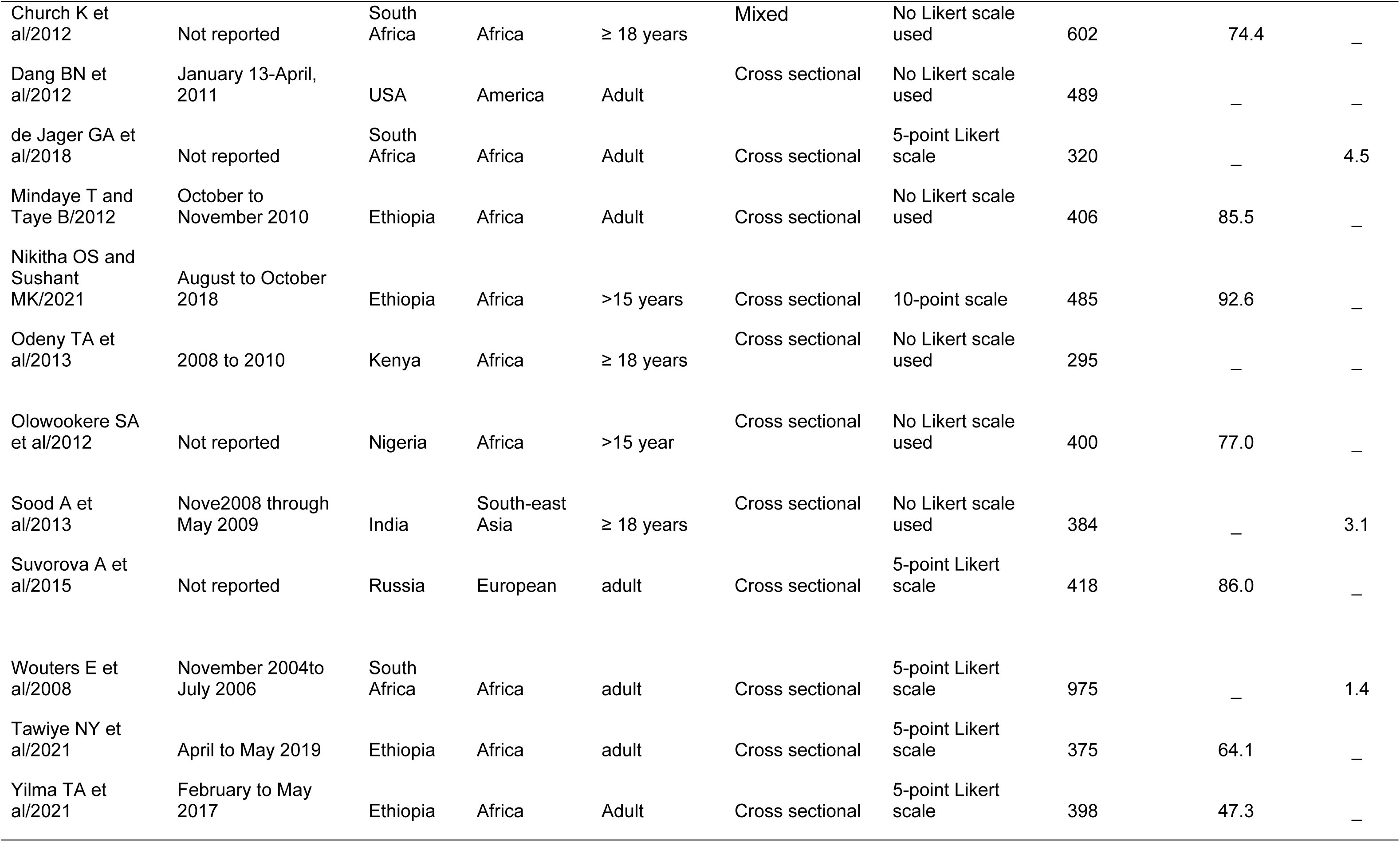
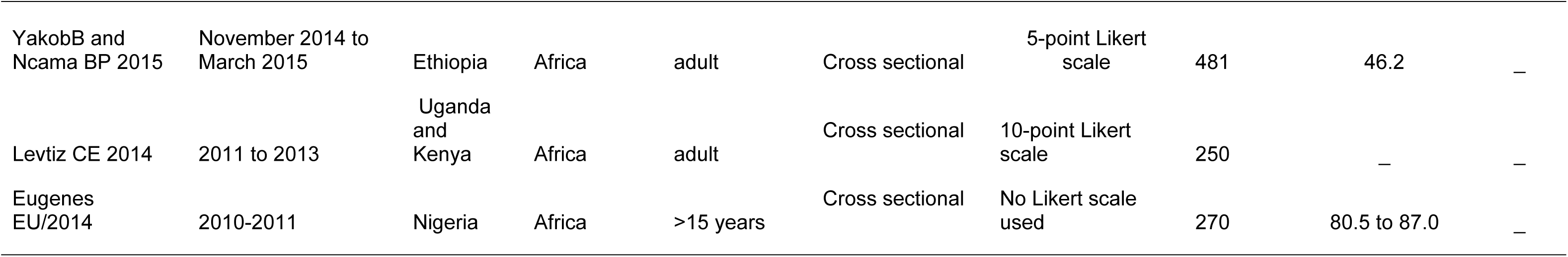
Characteristics of included articles

### Dimensions in the satisfaction tool

Thirty-one dimensions were used in the satisfaction measurement tool. Fifteen were used at once which are assurance, empathy, managing therapy, healthcare provider understood clients’ needs, management of staff, clients’ decisions in treatment, benefited more than expected, discrimination and grievance redressal, support, organisation of the services, perceived improved health, perception and knowledge of clients on ARV drugs, always obtained medication and prompt attention. Figure two depicts the dimensions shared by two or more articles (Figure 2).

**Figure 2:**
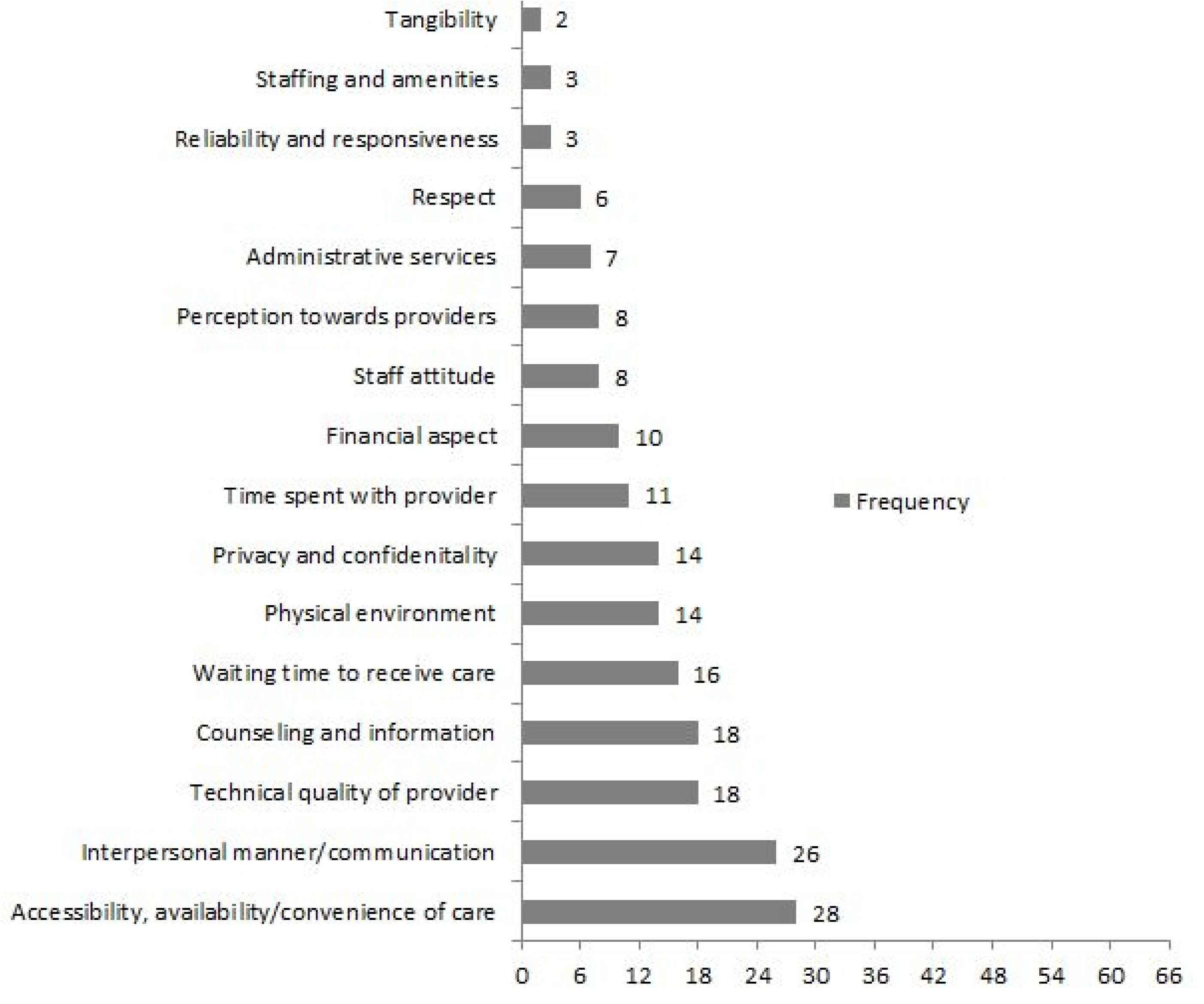
Dimensions employed in the assessment of satisfaction

### Satisfaction

Forty-two papers have appeared with percentage of satisfaction, while the percentage of dis-satisfied patients was published in two studies. In Ethiopia, fourteen studies found percentages of satisfaction ranging from 46.2% to 92.6%. In Nigeria, nine studies were published with reported percentages of satisfied HIV clients ranging from 56.5% to 99.6%: South Africa (between 65 and 98.0%), India (61.3% and 88.0% overall percentage), Brazil (80.0%), Tanzania (70.0 and 92.3%), Vietnam (42.4%), Cameroon (91.2%), France (16.0% and 15.9% for dimension), USA (85.5%), Canada (88.0%), Russia (86.0%), and Zambia (74.1%) (Table 1).

### Determinants of satisfaction of People living with HIV

Thirty-four articles have reported on determinants of satisfaction. There were healthcare settings (health service and health worker-related) and patient-related determinants.

#### Health service factors

Accessibility, authority/health facility level, healthcare cost and HIV care integration to primary healthcare are health service-related factors.

Perceived convenience of the location of health centre (75), perceived convenience and cleanliness (47, 61, 72), perceived availability of the services (50), higher health system responsiveness score (19), better perceptions of resources and services (37), perceiving the high number of nurses working at the treatment centre (64), presence of drinking water in the health facility (72), availability of ordered laboratory tests (56), visiting the health centre less frequently (20), HIV clients who travel distance that took short time (18, 57, 63), those disagreed with the access of toilets than strongly disagree (46) had a positive association with satisfaction of people living with HIV. The absence of signs and directions to ART clinics reduced client satisfaction (63)

Public-owned healthcare facility users (45) were less satisfied than the private-owned facility. In contrast, those caregivers whose HIV-infected children have received care in private facilities (49) and HIV clients attending rural facilities (37) were less satisfied. Patients who got services in the decentralised than centralised health facility (75), tertiary healthcare facility than secondary health facility (41), health centre (19), dispensary level than a hospital (31), club than clinic (35), and did not possess health care cards (20) were more satisfied. HIV clients who attended provincial and district clinics than central clinics (83, 84) and costs over 120 birrs (54) were less satisfied. HIV service integration with primary healthcare increases satisfaction (29).

#### Health worker factors

Health worker determinants were technical competency, waiting time to receive care, time to advise or consult clients, information provision, interpersonal relationship, knowledge and attitude of healthcare provider, and confidentiality and privacy. Technical competency encompasses healthcare service adherence to the standard of care, or competency, ability and performance of healthcare workers by following the professional code of ethics. Information provision is offering information and health education about drug-drug and drug-food interactions and guidance towards treatment. Interpersonal relationship refers to therapeutic communication in the way of sharing information, active listening, participation of patients, reflecting of acceptable behaviour and patient-centre interactions. Confidentiality in healthcare incorporates keeping or not disclose information and medical records of patients to a third person.

Those who perceived receiving a high quality of care (19, 47, 78), received services from nurses and health officers than doctors (54), waited a long time to receive care (20, 29, 33, 43, 56, 61, 72, 75), have got advice long time (51, 53), received information or health education (29, 43, 46), good interpersonal relationship (46, 50), perceived received care from knowledgeable and good attitudes/behaviour healthcare providers (20, 37, 45, 72, 82), perceived confidentiality is maintained (20, 45, 50, 72), perceived privacy during receiving services (46, 50), perceived low-level than high-level stigma and discrimination (51, 57), and perceived respected by nurses (75) were more likely to be satisfied.

One study revealed HIV clients were dissatisfied with the information provided during specimen collection (47).

#### Patient (sociodemographic and health condition) determinants

Age, gender, residency, marital status, religion, educational status, employment status, monthly income, patient support, health status, being on HAART, adherence to care, treatment duration, HIV disclosure status and being a drugs user were identified as unique modifiable and non-modifiable personal determinants.

Older age groups (18, 21, 44, 46, 57, 63) and female HIV clients (28, 29, 44, 61, 64) were more likely to be satisfied. In another two studies, females were less satisfied (45, 82). Two studies found that urban residency was more likely to be satisfied with HIV care services (44, 45). Conversely, another study revealed those who were living in rural areas were more likely to be satisfied (65). Those who married (44, 46) and divorced (46) than single, and those living with spouses or partners were more satisfied (84). Five studies determined the more educated were positively associated with satisfaction (28, 44, 50, 53, 57). On the other hand, four articles revealed the reverse in which less-educated HIV patients were more satisfied (18, 65, 82, 83). Those employed were more satisfied based on three studies (20, 44, 64). Other studies explained that unemployed patients (31), merchants (60), daily laborers (46), farmers (46) and students (60) were more satisfied than a government employee. In contrast, another study revealed that daily laborers were less satisfied than a merchant (51).

Those who had no monthly income were less satisfied (46), where as another study identified that the richest HIV clients were less satisfied (84).

Patients who perceived have support from friends and one’s family and comfortable housing conditions were more satisfied (21, 51, 81).

Patients who were perceived to have impaired mental health (19, 78), those on WHO stage II, III and IV (83), and patients having problems of pain or discomfort (83) had lower satisfaction level. Those who had hepatitis C co-infection (21), cluster of differentiation 4 (CD4) count >500 (53), being on HAART (78), taking treatment for 2 years and above (53), had fewer HAART side effects (21), having good adherence to HIV care (18, 65), those disclosed their seropositivity (50) and rated their disclosure status as very good (51) had more level of satisfaction. Another study revealed those who had higher CD4 counts (84) and drug users (84) were less satisfied.

## Discussion

The overall HIV/AIDS client satisfaction level has appeared inconclusive due to different settings and health systems, and differences in measurement tools. Based on this review, the majority of articles included accessibility and availability dimensions in the measurement tool of satisfaction. It is difficult to conclude that overall satisfaction is higher or lower even in countries with ineffective treatment outcomes (86), unfair distribution of HIV and general healthcare resources (87), ineffective financial risk protection (88), low adherence to clinical practice guidelines, poor service delivery indicators, suboptimal clinical practice (89–93), the application of emerging medical technologies to care (94, 95), an increasing discriminatory attitude (96), and a low progress record in ending the AIDS pandemic (97, 98).

With these challenges, findings in specific WHO regions were apparent. In the Americas region, a large percentage of people living with HIV are satisfied with healthcare services. In line with this, the majority of patients other than those with HIV in the USA were highly satisfied with the services received at outpatient medical services (99). In Canada, the majority of clients come to healthcare for needs other than HIV-related services grade the hospital services as very good (100). In contrast to the American region, people living with HIV make up less than half of study participants satisfaction with healthcare services in Vietnam. In contrast, clients who received care other than HIV-related services reported a higher satisfaction level in the same country (101). In the European region, two extremes were reported: the lowest in France and the highest in Russia. Another study in Russia showed that clients who received other healthcare services claimed a lower rate of satisfaction with the quality of care (102). In the southeast Asia region, the percentage of satisfied people living with HIV were almost comparable with TB (103) and type-2 diabetes mellitus patients (104).

This review identified the satisfaction of people living with HIV associated with healthcare settings, health workers, and patient-related issues. A global review of literature about patient satisfaction other than HIV clients revealed similar findings to this review (105). Another review from developing countries about maternal care revealed that healthcare setting, health service factors, socio-economic status, and health status affect the satisfaction level of clients (106). Evidence from a study in Pakistan also identified personal and health worker-related variables associated with the satisfaction of diabetes mellitus patients (107).

To see individual determinants, accessibility, authority/health facility level, healthcare cost, and HIV care integration into primary healthcare were health service-related factors. Accessibility due to unavailability of services, locations and distances of health facilities, financial problems, individual acceptability of the service, and waiting much time to receive healthcare (108–110) create unmet healthcare needs. HIV integration services with primary healthcare increase patient satisfaction by minimising waiting time and enhancing coordination of care (111). Integrated HIV services at the primary healthcare level default to reaching most of the community without going a long distance, and an issue of equity, accessibility, and availability may be resolved as a result.

Health worker-related determinants were technical competency, waiting time to receive care, time to advise or consult clients, information provision, interpersonal relationships, knowledge and attitude of healthcare providers, stigma and discrimination, confidentiality, and privacy. Technical competence is part of general competence and the main focus of patient perception, mainly about the skill of providers (112). Patients’ perception of the positive attitude and behaviour of care providers could arise from the caring, respectful, and friendly approach of healthcare providers which affect satisfaction (113). Consultation time was directly associated with HIV patients’ satisfaction. Due to the chronic nature of lifelong treatment or non-curable disease, HIV patients might be stressed, anxious, and have an impaired quality of life (114–116). Those who received advice and counselling, had good communication with care giver, felt privacy and confidentially kept could be more satisfied.

Clients’ demographic conditions influenced the way they feel to the services they received. Except age and gender, which need age and gender-sensitive services, others were modifiable determinants. This implies due to the unique features of HIV and longevity, provision of HIV/AIDS services should consider comprehensive parameters. Therefore, culture and context specific services should also be implemented that should cover both social determinants of health and social determinants process.

### Limitation

Articles other than English were excluded. The pooled global estimation, pooled estimated variation across the regions, and trend over time of satisfaction were not explored because of the variety of measurement tools used across studies.

## Conclusions

Implementing multi-pronged interventions in healthcare settings and patient-related factors could foster client satisfaction with health services. It is vital to improve HIV care integration at the primary healthcare level, establish or strengthen the financing of HIV care, digitalise healthcare settings, invest in pre-service education and in-service training, provide culturally sensitive services, establish or strengthen and coordinate social work with the health system, strengthen behavioural change communication and rehabilitative services, and enhance political commitment and institutional capacity. These can serve as the building blocks of a framework for action to improve the determinants of HIV client’s satisfaction.

## Data Availability

All relevant data are within the manuscript and its Supporting Information files.

## Acknowledgement

We acknowledge Mr. Scott Macintyre, Librarian at The University of Queensland, Australia for assisting search of MeSH terms and building search strings and The University of Queensland, Australia for allowed to use databases which are not publicly available.

## Authors’ contribution

AE: conceptualization, identify research questions, built search strategy, literature searching, write manuscript draft and final report; YA: conceptualisation, identify research questions, built search strategy, and supervision; CFG: conceptualisation, and supervision; FA: supervision and revision; MDC: revision and

## Fund

Authors have not received any fund to conduct this research.

## Supplementary files

Supplementary file 1: Search strategies

Supplementary file 2: Quality status of included articles

## References

1. UNAIDS. Preliminary UNAIDS 2021 epidemiological estimates: GLOBAL HIV STATISTICS : https://www.unaids.org/sites/default/files/media_asset/UNAIDS_FactSheet_en.pdf; Access Date: 17 March 2021. 2021.

2. UNAIDS. 90-90-90 An ambitious treatment target to help end the AIDS epidemic. Geneva, Switzerland; 2014.

3. United Nations. Transforming our world: The 2030 agenda for sustainable development. New York: United Nations, Department of Economic and Social Affairs. 2015.

4. World Health Organization. Making fair choices on the path to universal health coverage: Final report of the WHO Consultative Group on Equity and Universal Health Coverage. 2014.

5. World Health Organization. World Health Assembly:Sustainable health financing, universal coverage and social health insurance. Fifty-eighth World Health Assembly edition Geneva: World Health Organization. 2005.

6. World Health Organization. Manual on use of routine data quality assessment (RDQA) tool for TB monitoring. World Health Organization, 2011 9241501243.

7. Dang BN, Westbrook RA, Black WC, Rodriguez-Barradas MC, Giordano TP. Examining the link between patient satisfaction and adherence to HIV care: a structural equation model. PloS one. 2013;8(1):e54729.

8. Wahyuhadi J, Hidayah N, Aini Q. Remuneration, Job Satisfaction, and Performance of Health Workers During the COVID-19 Pandemic Period at the Dr. Soetomo Hospital Surabaya, Indonesia. Psychology Research and Behavior Management. 2023:701–11.

9. Dwivedi R, Anand MS. Interrelating Employee Satisfaction & Customer Satisfaction in Healthcare Industry. International Bulletin of Management and Economics. 2019;10:214–32.

10. Grusky O, Marelich WD, Erger J, Mann T, Roberts KJ, Steers WN, et al. Evaluation of a brief low-cost intervention to improve antiretroviral treatment decisions. AIDS Care - Psychological and Socio-Medical Aspects of AIDS/HIV. 2003;15(5):681–7.

11. Kebede H, Tsehay T, Necho M, Zenebe Y. Patient satisfaction towards outpatient pharmacy services and associated factors at dessie town public hospitals, south Wollo, north-east Ethiopia. Patient Preference and Adherence. 2021;15:87–97.

12. Wong E, Mavondo F, Fisher J. Patient feedback to improve quality of patient-centred care in public hospitals: a systematic review of the evidence. BMC health services research. 2020;20:1–17.

13. Kwesiga D, Kiwanuka S, Kiwanuka N, Mafigiri D, Kakande N. The clients’ voice: satisfaction with HIV/AIDS care in a public and private health facility in Kabale District, Uganda. Value in Health. 2014;17(3):A140.

14. Dapaah JM, Senah KA. HIV/AIDS clients, privacy and confidentiality; the case of two health centres in the Ashanti Region of Ghana. BMC medical ethics. 2016;17:1–10.

15. Ford RC, Bach SA, Fottler MD. Methods of measuring patient satisfaction in health care organizations. Health care management review. 1997:74–89.

16. Banda S, Nkungula N, Chiumia IK, Rylance J, Limbani F. Tools for measuring client experiences and satisfaction with healthcare in low-and middle-income countries: a systematic review of measurement properties. BMC Health Services Research. 2023;23(1):133.

17. Linder-Pelz S. Toward a theory of patient satisfaction. Social science & medicine. 1982;16(5):577–82.

18. Dixit S, Verma N, Shrivastava N, Sharma M, Pradhan SK, Agarwal S. Patient satisfaction with ART centre services among people living with HIV: a cross sectional study in a tertiary care hospital, Chhattisgarh, India. Int J Community Med Public Health [Internet]. 2018;5(6):2564–71.

19. Yakob B, Ncama BP. Client satisfaction: Correlates and implications for improving HIV/AIDS treatment and care services in southern Ethiopia. International Health. 2016;8(4):292–8.

20. Chow MYK, Li M, Quine S. Client satisfaction and unmet needs assessment: Evaluation of an HIV ambulatory health care facility in Sydney, Australia. Asia-Pacific Journal of Public Health. 2012;24(2):406–14.

21. Préau M, Protopopescu C, Raffi F, Rey D, Chêne G, Marcellin F, et al. Satisfaction with care in HIV-infected patients treated with long-term follow-up antiretroviral therapy: The role of social vulnerability. AIDS Care - Psychological and Socio-Medical Aspects of AIDS/HIV. 2012;24(4):434–43.

22. Okafoagu N, Ango J, Gana G, Kaoje A, Raji M, Oche M, et al. Clients satisfaction with anti retroviral therapy services in a tertiary hospital in Sokoto, Nigeria. Journal of AIDS and HIV Research. 2013;5(9):328–33.

23. Chimbindi N, Bärnighausen T, Newell ML. Patient satisfaction with HIV and TB treatment in a public programme in rural KwaZulu-Natal: evidence from patient-exit interviews. BMC Health Serv Res. 2014;14:32.

24. Page MJ MJ, Bossuyt PM, Boutron I, Hoffmann TC, Mulrow CD, et al.,. The PRISMA 2020 statement: an updated guideline for reporting systematic reviews. BMJ 2021;372:n71. doi: 10.1136/bmj.n71.

25. Bramer WM, Giustini D, de Jonge GB, Holland L, Bekhuis T. De-duplication of database search results for systematic reviews in EndNote. Journal of the Medical Library Association: JMLA. 2016;104(3):240.

26. Modesti PA, Reboldi G, Cappuccio FP, Agyemang C, Remuzzi G, Rapi S, et al. Panethnic differences in blood pressure in Europe: a systematic review and meta-analysis. PloS one. 2016;11(1):e0147601.

27. Stein MD, Fleishman J, Mor V, Dresser M. FACTORS ASSOCIATED WITH PATIENT SATISFACTION AMONG SYMPTOMATIC HIV-INFECTED PERSONS. Medical Care. 1993;31(2):182–8.

28. Paul NI, Ugwu RO. Caregivers/Patients Perception and Satisfaction with Outpatient HIV Services at the University of Port Harcourt Teaching Hospital (UPTH), Port Harcourt, Nigeria. Journal of Scientific Research and Reports. 2019:1–9.

29. Odeny TA, Penner J, Lewis-Kulzer J, Leslie HH, Shade SB, Adero W, et al. Integration of HIV care with primary health care services: Effect on patient satisfaction and stigma in Rural Kenya. AIDS Research and Treatment. 2013;2013.

30. Church K, Wringe A, Fakudze P, Kikuvi J, Simelane D, Mayhew SH. The relationship between service integration and client satisfaction: A mixed methods case study within HIV services in a high prevalence setting in Africa. AIDS Patient Care and STDs. 2012;26(11):662–73.

31. Ndayongeje J, Kazaura M. Satisfaction of patients attending public HIV or AIDS care and treatment centers in Kinondoni district, Tanzania. International Quarterly of Community Health Education. 2017;37(2):113–9.

32. Magoro M, Hoque M, Van Der Heever H. ART patients’ satisfaction level regarding comprehensive HIV and AIDS care management and antiretroviral treatment programme in Pretoria. Southern African Journal of Epidemiology and Infection. 2012;27(2):71–5.

33. Karunamoorthi K, Rajalakshmi M, Babu SM, Yohannes A. HIV/AIDS patient’s satisfactory and their expectations with pharmacy service at specialist antiretroviral therapy (ART) units. Eur Rev Med Pharmacol Sci. 2009;13(5):331–9.

34. Wouters E, Heunis C, van Rensburg D, Meulemans H. Patient satisfaction with antiretroviral services at primary health-care facilities in the Free State, South Africa - a two-year study using four waves of cross-sectional data. Bmc Health Services Research. 2008;8.

35. de Jager GA, Crowley T, Esterhuizen TM. Patient satisfaction and treatment adherence of stable human immunodeficiency virus-positive patients in antiretroviral adherence clubs and clinics. African Journal of Primary Health Care and Family Medicine. 2018;10(1).

36. Osungbade KO, Shaahu VN, Owoaje EE, Adedokun BO. Patients’ satisfaction with quality of anti-retroviral services in Central Nigeria: implications for strengthening private health services. World Journal of Preventive Medicine. 2013;1(3):11–8.

37. Dansereau E, Masiye F, Gakidou E, Masters SH, Burstein R, Kumar S. Patient satisfaction and perceived quality of care: Evidence from a cross-sectional national exit survey of HIV and non-HIV service users in Zambia. BMJ Open. 2015;5(12).

38. Olanrewaju OP, Oyindamola AO, Ajidat JA, Oluyemi J, Ayotunde OO, Osagie OMA, et al. Patients’ Treatment Satisfaction: Experiences of HIV Patients in a Tertiary Hospital in Southwest Nigeria. Science Journal of Clinical Medicine. 2020;9(3):50.

39. Okoye MO, Ukwe VC, Okoye TC, Adibe MO, Ekwunife OI. Satisfaction of HIV patients with pharmaceutical services in South Eastern Nigerian hospitals. International Journal of Clinical Pharmacy. 2014;36(5):914–21.

40. Getenet H, Haileamlak A, Tegegn A. CLIENTS’SATISFACTION WITH ANTI RETROVIRAL THERAPY SERVICES AT JIMMA UNIVERSITY SPECIALIZED HOSPITAL. Ethiopian Journal of Health Sciences. 2008;18(2).

41. Nwabueze S, Adogu P, Ilika A, Asuzu M. Comparative analysis of patient satisfaction levels in HIV/AIDS care in secondary and tertiary health care facilities in Nigeria. Afrimedic Journal. 2010;1(2):1–9.

42. Abebe TB, Erku DA, Gebresillassie BM, Haile KT, Mekuria AB. Expectation and satisfaction of HIV/AIDS patients toward the pharmaceutical care provided at Gondar university referral hospital, northwestern Ethiopia: A cross-sectional study. Patient Preference and Adherence. 2016;10:2073-82.

43. Girmay A, Tilahun Z, Akele MZ. Adult HIV/AIDS Patients’ Level of Satisfaction on Pharmaceutical Service: Institutional Prospective Cross Sectional Study. 2019.

44. Azuike E, Adinma E, Umeh U, Njelita I, Anemeje O, Aniemena R. CLIENTS’SATISFACTION WITH WAITING TIME IN HIV TREATMENT CENTRES: AN URBAN RURAL COMPARISON IN ANAMBRA STATE, NIGERIA. Medico Research Chronicles. 2017;4(01):61–76.

45. Anosike A, Olakunde BO, Adeyinka DA, Ezeokafor C, Amanze O, Mathews O, et al. Clients’ satisfaction with HIV treatment and care services in Nigeria. Public Health. 2019;167:50–4.

46. Atsebeha KG, Chercos DH. High antiretroviral therapy service delivery satisfaction and its’ associated factors at Midre-genet hospital; Northwest Tigray, Ethiopia. BMC Health Services Research. 2018;18(1).

47. Belay M, Abrar S, Bekele D, Daka D, Derbe M, Birhaneselassie M. HIV⁄ AIDS Patients’ satisfaction on ART laboratory service in selected governmental hospitals, Sidamma Zone, southern Ethiopia. Sci J Public Health. 2013;1:85.

48. Bezuidenhout S, Ogunsanwo DA, Helberg EA. Patient satisfaction at accredited antiretroviral treatment sites in the Gert Sibande District. African journal of primary health care & family medicine. 2014;6(1):E1–6.

49. Chamla D, Asadu C, Adejuyigbe E, Davies A, Ugochukwu E, Umar L, et al. Caregiver satisfaction with paediatric HIV treatment and care in Nigeria and equity implications for children living with HIV. AIDS Care - Psychological and Socio-Medical Aspects of AIDS/HIV. 2016;28:153–60.

50. Doyore F, Moges B. Client satisfaction to antiretroviral treatment services and associated factors among clients attending ART clinics in Hossana town, southern Ethiopia. Clin Res. 2016;2(6):6.

51. Habtamu A, Kifle Y, Ejigu Y. Client Satisfaction and its Determinants with Anti-Retroviral Therapy (ART) Services in Public Hospitals of West Wollega Zone, Ethiopia: a Cross Sectional Study. Galore Int J Appl Sci Humanit. 2017;1:1–16.

52. Kagashe GAB, Rwebangila F. Patient satisfaction with health care services provided at HIV clinics at Amana and Muhimbili hospitals in Dar Es Salaam. African Health Sciences. 2011;11(SPEC. ISSUE):S60–S6.

53. Nigussie T, Aferu T, Mamo Y, Feyisa M. Patient satisfaction with hiv and aids services in mizan-tepi university teaching hospital, southwest ethiopia. HIV/AIDS - Research and Palliative Care. 2020;12:403–10.

54. Asfaw E, Dominis S, Palen JGH, Wong W, Bekele A, Kebede A, et al. Patient satisfaction with task shifting of antiretroviral services in Ethiopia: Implications for universal health coverage. Health Policy and Planning. 2014;29:ii50–ii8.

55. Buluba SE, Mawi NE, Tarimo EAM. Clients’ satisfaction with HIV care and treatment centres in Dar es Salaam, Tanzania: A cross-sectional study. PLoS One. 2021;16(2):e0247421.

56. Mindaye T, Taye B. Patients satisfaction with laboratory services at antiretroviral therapy clinics in public hospitals, Addis Ababa, Ethiopia. BMC research notes. 2012;5:184.

57. Yimer Tawiye N, Mekonnen Assefa Z, Gizeyatu Zengye A. Patient satisfaction and associated factors among adults attending ART clinic at Dessie refferal Hospital, Amhara Region, Ethiopia. International Journal of Africa Nursing Sciences. 2021;14.

58. Olowookere SA, Fatiregun AA, Ladipo MMA, Akenova YA. Reducing waiting time at a Nigerian HIV treatment clinic: Opinions from and the satisfaction of people living with HIV/AIDS. Journal of the International Association of Physicians in AIDS Care. 2012;11(3):188–91.

59. Yakob B, Ncama BP. Perceived quality of HIV treatment and care services in Wolaita Zone of southern Ethiopia: A cross-sectional study. BMJ Open. 2015;5(12).

60. Yilma TA, Beedemariam Gebretekle G, Gedif Fenta T. Patient Satisfaction with HIV/AIDS Services in Health Centers of East Shoa Zone, Oromia, Ethiopia: A Cross-Sectional Study. Health Services Insights. 2021;14:11786329211003106.

61. Levitz CE. Effect of patient satisfaction with a health care facility on HIV outcomes: A facility-level study in eastern Africa 2014.

62. Mukamba N, Chilyabanyama ON, Beres LK, Simbeza S, Sikombe K, Padian N, et al. Patients’ Satisfaction with HIV Care Providers in Public Health Facilities in Lusaka: A Study of Patients who were Lost-to-Follow-Up from HIV Care and Treatment. AIDS Behav. 2020;24(4):1151–60.

63. Adissu G, Biks GA, Tamirat KS. Patient satisfaction with antiretroviral therapy services and associated factors at Gondar town health centers, Northwest Ethiopia: An institution-based cross-sectional study. BMC Health Services Research. 2020;20(1).

64. Wung BA, Peter NF, Atashili J. Clients’ satisfaction with HIV treatment services in Bamenda, Cameroon: A cross-sectional study. BMC Health Services Research. 2016;16(1).

65. Umeokonkwo CD, Aniebue PN, Onoka CA, Agu AP, Sufiyan MB, Ogbonnaya L. Patients’ satisfaction with HIV and AIDS care in Anambra State, Nigeria. PLoS One. 2018;13(10):e0206499.

66. Eugenes EU. CLIENTS’ SATISFACTION WITH SERVICES FOR CARE, SUPPORT AND TREATMENT OF HIV/AIDS IN ASOKORO GENERAL HOSPITAL, ABUJA. 2014.

67. Shukla M. A Cross Sectional Study to Assess The Patient’s Satisfaction With Services Provided At Antiretroviral Therapy Centers in Lucknow. Medical Science. 2016;5(5).

68. Aung MN, Moolphate S, Kitajima T, Siriwarothai Y, Takamtha P, Katanyoo C, et al. Satisfaction of HIV patients with task-shifted primary care service versus routine hospital service in northern Thailand. Journal of Infection in Developing Countries. 2015;9(12):1360–6.

69. Sunita D, Gupta A, Mahajan A, Sachdeva A. A cross sectional study to assess the satisfaction level among people living with HIV/AIDS in a hilly state of northern India. International Journal of Community Medicine and Public Health. 2018;5(6):2373.

70. Vahab SA, Madi D, Ramapuram J, Bhaskaran U, Achappa B. Level of Satisfaction Among People Living with HIV (PLHIV) Attending the HIV Clinic of Tertiary Care Center in Southern India. Journal of Clinical and Diagnostic Research. 2016;10(4):OC8–OC10.

71. Chander V, Bhardwaj AK, Raina SK, Bansal P, Agnihotri RK. Scoring the medical outcomes among HIV / AIDS patients attending antiretroviral therapy center at Zonal Hospital, Hamirpur, using patient satisfaction questionnaire (PSQ-18). Indian Journal of Sexually Transmitted Diseases. 2011;32(1):19–22.

72. Devnani M, Gupta AK, Wanchu A, Sharma RK. Factors associated with health service satisfaction among people living with HIV/AIDS: A cross sectional study at ART center in Chandigarh, India. AIDS Care - Psychological and Socio-Medical Aspects of AIDS/HIV. 2012;24(1):100–7.

73. Sood A, Mazta S, Sharma A, Bhardwaj A, Raina SK, Chander V. Scoring satisfaction among patients, attending ART Centre of a medical college in north-west India. AIDS Care - Psychological and Socio-Medical Aspects of AIDS/HIV. 2013;25(12):1477–80.

74. Nikitha OS, Sushant MK. Client satisfaction of antiretroviral therapy service delivery: A cross-sectional study at an antiretroviral therapy center. International Journal of Applied and Basic Medical Research. 2021;11(1):14–20.

75. Leon C, Koosed T, Philibert B, Raposo C, Benzaken AS. HIV/AIDS health services in Manaus, Brazil: Patient perception of quality and its influence on adherence to antiretroviral treatment. BMC Health Services Research. 2019;19(1).

76. Burke JK, Cook JA, Cohen MH, Wilson T, Anastos K, Young M, et al. Dissatisfaction with medical care among women with HIV: Dimensions and associated factors. AIDS Care - Psychological and Socio-Medical Aspects of AIDS/HIV. 2003;15(4):451–62.

77. Dang BN, Westbrook RA, Rodriguez-Barradas MC, Giordano TP. Identifying Drivers of Overall Satisfaction in Patients Receiving HIV Primary Care: A Cross-Sectional Study. Plos One. 2012;7(8).

78. Burke-Miller JK, Cook JA, Cohen MH, Hessol NA, Wilson TE, Richardson JL, et al. Longitudinal relationships between use of highly active antiretroviral therapy and satisfaction with care among women living with HIV/AIDS. American Journal of Public Health. 2006;96(6):1044–51.

79. Guilherme JA, Yamaguchi MU, Massuda EM. HIV/AIDS PATIENTS SATISFACTION WITH THE SPECIALIZED CARE SERVICE. Rev Min Enferm. 2019.

80. Marshall BC, Cunny KA, Lawson KA. HIV-positive males’ satisfaction with pharmacy services. Journal of the American Pharmaceutical Association. 1997;37(1):66–75.

81. Tsasis P, Tsoukas C, Deutsch G. Evaluation of patient satisfaction in a specialized HIV/AIDS care unit of a major hospital. AIDS Patient Care and STDs. 2000;14(7):347–9.

82. Li L, Comulada WS, Wu Z, Ding Y, Zhu W. Providers’ HIV-related avoidance attitude and patient satisfaction. Health Expectations. 2013;16(1):105–12.

83. Tran BX, Dang AK, Vu GT, Tran TT, Latkin CA, Ho CSH, et al. Patient satisfaction with HIV services in Vietnam: Status, service models and association with treatment outcome. Plos One. 2019;14(11).

84. Tran BX, Nguyen NP. Patient satisfaction with HIV/AIDS care and treatment in the decentralization of services delivery in Vietnam. PLoS One. 2012;7(10):e46680.

85. Suvorova A, Belyakov A, Makhamatova A, Ustinov A, Levina O, Tulupyev A, et al. Comparison of satisfaction with care between two different models of HIV care delivery in St. Petersburg, Russia. Aids Care-Psychological and Socio-Medical Aspects of Aids/Hiv. 2015;27(10):1309–16.

86. Akachi Y, Kruk ME. Quality of care: measuring a neglected driver of improved health. Bull World Health Organ. 2017;95(6):465–72.

87. Sinha G, Peters DH, Bollinger RC. Strategies for gender-equitable HIV services in rural India. Health policy and planning. 2009;24(3):197–208.

88. Assebe LF, Negussie EK, Jbaily A, Tolla MTT, Johansson KA. Financial burden of HIV and TB among patients in Ethiopia: a cross-sectional survey. BMJ open. 2020;10(6):e036892.

89. Kruk ME, Leslie HH, Verguet S, Mbaruku GM, Adanu RM, Langer A. Quality of basic maternal care functions in health facilities of five African countries: an analysis of national health system surveys. The lancet global health. 2016;4(11):e845–e55.

90. Martin GH, Pimhidzai O. Service delivery indicators: Kenya. 2013.

91. Peabody JW, Tozija F, Munoz JA, Nordyke RJ, Luck J. Using vignettes to compare the quality of clinical care variation in economically divergent countries. Health services research. 2004;39(6p2):1951–70.

92. Das J, Hammer J, Leonard K. The quality of medical advice in low-income countries. Journal of Economic perspectives. 2008;22(2):93–114.

93. Graversen L, Christensen B, Borch-Johnsen K, Lauritzen T, Sandbaek A. General practitioners’ adherence to guidelines on management of dyslipidaemia: ADDITION-Denmark. Scandinavian journal of primary health care. 2010;28(1):47–54.

94. Le D-N, Van Le C, Tromp JG, Nguyen GN. Emerging technologies for health and medicine: virtual reality, augmented reality, artificial intelligence, internet of things, robotics, industry 4.0: John Wiley & Sons; 2018.

95. DeMaio J, Schwartz L, Cooley P, Tice A. The application of telemedicine technology to a directly observed therapy program for tuberculosis: A pilot project. Clinical Infectious Diseases. 2001;33(12):2082–4.

96. UNAIDS Global AIDS update. Seizing the moment: tackling entrenched inequalities to end epidemics. 2020.

97. Chakaya J, Khan M, Ntoumi F, Aklillu E, Fatima R, Mwaba P, et al. Global Tuberculosis Report 2020–Reflections on the Global TB burden, treatment and prevention efforts. International Journal of Infectious Diseases. 2021.

98. Assefa Y, Gilks CF. Ending the epidemic of HIV/AIDS by 2030: Will there be an endgame to HIV, or an endemic HIV requiring an integrated health systems response in many countries? International Journal of Infectious Diseases. 2020;100:273–7.

99. Tehrani AB, Feldman SR, Camacho FT, Balkrishnan R. Patient satisfaction with outpatient medical care in the United States. Health Outcomes Research in Medicine. 2011;2(4):e197–e202.

100. lanie J, e Davidson JLSMRKKH, Naomi D. Patient Experiences in Canadian Hospitals. Healthcare Quarterly. 2019;22(3):12–4.

101. Quyen BTT, Ha NT, Van Minh H. Outpatient satisfaction with primary health care services in Vietnam: Multilevel analysis results from The Vietnam Health Facilities Assessment 2015. Health Psychology Open. 2021;8(1):20551029211015117.

102. Statista Research Department. Level of satisfaction of the French regarding the quality of medical care in 2016. (https://www.statista.com/statistics/767000/satisfaction-quality-medical-care-french/) Access date 21 July 2021. France; 2016. Contract No.: 21.

103. Rai N, Singh S, Kushwah S, Dubey D. A cross sectional study on evaluation of satisfaction level of TB patients enrolled for directly observed treatment, short course chemotherapy in a district of Central India. Int J Community Med Public Heal. 2017;4(1):5–8.

104. Priya T, Jayaseelan V, Krishnamoorthy Y, Sakthivel M, Majella MG. Patient’s Experiences and Satisfaction in Diabetes Care and Out-of-Pocket Expenditure for Follow-Up Care Among Diabetes Patients in Urban Puducherry, South India. Journal of Patient Experience. 2020;7(6):1445–9.

105. Batbaatar E, Dorjdagva J, Luvsannyam A, Savino MM, Amenta P. Determinants of patient satisfaction: a systematic review. Perspectives in public health. 2017;137(2):89–101.

106. Srivastava A, Avan BI, Rajbangshi P, Bhattacharyya S. Determinants of women’s satisfaction with maternal health care: a review of literature from developing countries. BMC pregnancy and childbirth. 2015;15(1):1–12.

107. Jalil A, Zakar R, Zakar MZ. Satisfaction of diabetes patients in public outpatient department: prevalance and determinants. Rawal Medical Journal. 2018;43(1):8–13.

108. Aday LA, Andersen R. A framework for the study of access to medical care. Health services research. 1974;9(3):208.

109. Chen J, Hou F. Unmet needs for health care. Health Rep. 2002;13(2):23–34.

110. Cavalieri M. Geographical variation of unmet medical needs in Italy: a multivariate logistic regression analysis. International journal of health geographics. 2013;12(1):1–11.

111. Bernard S, Tailor A, Jones P, Alexander DE. Addressing the medical and support service needs of people living with HIV (PLWH) through program collaboration and service integration (PCSI). Californian journal of health promotion. 2016;14(1):1.

112. Kak N, Burkhalter B, Cooper M-A. Measuring the competence of healthcare providers. Operations Research Issue Paper. 2001;2(1):1–28.

113. Halldorsdottir S. Caring and uncaring encounters in nursing and health care: Developing a theory: Linköpings universitet; 1996.

114. Peltzer K, Naidoo P, Matseke G, Louw J, Mchunu G, Tutshana B. Prevalence of psychological distress and associated factors in tuberculosis patients in public primary care clinics in South Africa. BMC psychiatry. 2012;12(1):1–9.

115. Duko B, Bedaso A, Ayano G. The prevalence of depression among patients with tuberculosis: a systematic review and meta-analysis. Annals of general psychiatry. 2020;19:1–11.

116. Belayneh Z, Mekuriaw B, Mehare T, Shumye S, Tsehay M. Magnitude and predictors of common mental disorder among people with HIV/AIDS in Ethiopia: a systematic review and meta-analysis. BMC public health. 2020;20:1–11.

